# The two costs of neglect: evaluating the health and economic impacts of anthrax in an endemic area of rural Africa

**DOI:** 10.1101/2025.06.03.25328853

**Authors:** O. Rhoda Aminu, Taya L. Forde, Mohammad Maksudur Rahman, Chobi Clement Chubwa, Loturiaki Korio, Nestory Mkenda, Deogratius Mshanga, Sabore Ole Moko, Sruti Pisharody, Matthew P. Rubach, Ruth N. Zadoks, Blandina T. Mmbaga, Roman Biek, Thomas L. Marsh, Tiziana Lembo

**Author notes:** Equal contributions (Thomas L. Marsh, Tiziana Lembo).

## Abstract

Endemic zoonoses have a dual burden, impacting human and animal health, as well as livelihoods due to livestock losses. One such disease is anthrax, caused by *Bacillus anthracis*. Data on the burden of anthrax on human and animal health are extremely limited, perpetuating neglect at the policy level in endemic areas and internationally. To address these data inadequacies, we quantified the widespread impacts of anthrax on highly affected communities, focusing on a hyper-endemic area of eastern Africa, the Ngorongoro Conservation Area (NCA) of northern Tanzania. Data were collected between 2016 and 2019 through cross-sectional household surveys, active surveillance, compilation of available hospital and human disease surveillance records, and willingness-to-pay choice experiments conducted within wider socioeconomic surveys. Human anthrax was reported by 34 (16%) of 209 households surveyed. Cases of anthrax in livestock were reported in 63 households (30%), and in all 20 households within Ngoile, the most affected area. The majority of sudden deaths in livestock (368 of 500 cases in 30 months of active surveillance) were confirmed to be caused by anthrax using microscopy or qPCR. Monetary losses due to livestock deaths were significant and during outbreaks commonly exceeded average monthly household income. Despite the dual burden, willingness to adopt anthrax mitigation measures was low. Likely due to resource limitations, households would only consider such measures if they had high efficacy, be it for animals or humans, but even then, price was still an important factor. We discuss the potential benefits that anthrax prevention could have on affected communities, and factors that might influence the success of possible interventions. This study demonstrates that in endemic settings where dependence on livestock productivity is high, the impacts of neglected zoonoses can be diverse, persistent and severe and highlights the challenge of identifying locally acceptable mitigation strategies.

## Introduction

Neglected zoonotic diseases (NZDs) are a subset of the neglected tropical diseases that can be transmitted between animals and people. These diseases have a dual burden: not only do they affect the health of both people and animals, but they also impact on people’s livelihoods through livestock deaths or decreased productivity [1]. The latter impacts are disproportionally felt by impoverished households [2,3] that depend on livestock for their livelihoods [4] as in most of sub-Saharan Africa [5]. Indeed, NZDs have a higher prevalence in marginalized and disadvantaged communities [6,7] where they continue to impact on health, wellbeing and economies, thereby perpetuating a cycle of poverty [8,9].

Quantitative assessments of the impacts of NZDs are critical to appropriately prioritize resources for intervention and reverse the cycle of neglect [7,10]. However, despite most of these diseases being notifiable, surveillance data are typically lacking because the most affected communities are remote, with poor infrastructure for effective monitoring of cases. For example, in rural Africa, infrastructure such as roads is often poor, surveillance and diagnostic capacity is limited for both the human and animal sectors, and health care services are thinly stretched [11]. Moreover, community use of health facilities is typically influenced by factors such as acceptability, affordability and geographic accessibility [12,13]. Thus, while large outbreaks affecting high numbers of animals and people [14–16] may be represented in official estimates, ongoing cases occurring outside of these major events are unlikely to present to a health facility and consequently go unreported. Because of this data gap, the burden of NZDs is grossly underestimated [10] and these diseases collectively receive less than 0.1% of international global health assistance [8].

In addition to direct health implications, NZDs have numerous financial ramifications such as costs associated with 1) livestock losses; 2) livestock treatment/vaccination; 3) medical treatment/vaccination, including direct costs, travel to medical facilities and time away from work; 4) surveillance and laboratory testing; and 5) other control measures (e.g., disposal of contaminated animal carcasses or products) [8]. Further societal impacts relate to the emotional stress or trauma related to animal and human morbidity and mortality [17], as well as impacts on food and asset security associated with livestock losses. To mitigate against these multiple impacts, both public and private investments are required [18]. However, due to under-resourced public health infrastructure and unreliable health coverage, in the most affected areas, private investment by individual households into livestock and household health is often necessary [18–20]. Consequently, poor livestock-income-dependent households are required to make complex choices on how best to allocate household resources, including investment towards animal and human health. Thus, understanding households’ willingness and ability to participate in disease mitigation interventions, amidst the broad range of competing priorities for expenditure, is integral to the evaluation of the likely success of alternative control strategies.

Among the major NZDs is anthrax, a serious disease in both humans and animals caused by the spore-forming bacterium *Bacillus anthracis*. It manifests as sudden deaths in herbivores (both livestock and wildlife), which become infected primarily through the ingestion of soil and/or plants contaminated by *B. anthracis* spores [21]. These spores can persist for several decades in the environment, remaining infectious even under extreme climatic conditions [22] and therefore representing a continuous health threat. Humans can become infected secondarily by handling or eating meat from contaminated carcasses [23], or coming into contact with contaminated soil or animal hides [24]. Human anthrax can take one of three main forms, depending on the route of exposure: cutaneous, gastrointestinal or inhalational. The latter two forms have high fatality rates if not treated with antibiotics, and the more common cutaneous anthrax cases can also develop systemic complications [25]. While human anthrax vaccines tend to be reserved for at-risk laboratory and military personnel, livestock vaccination is globally widespread and has been the mainstay of anthrax control in high-income countries [23]. Meanwhile, anthrax remains endemic in many low- and middle-income countries, including much of Africa (Fig 1). It is estimated that between 10 – 100 thousand cases of human anthrax occur worldwide each year [8], and that 1.8 billion people live in areas at risk of anthrax [26]. However, in endemic African settings, as with most NZDs, the impact of anthrax on the health and livelihoods of people and their livestock remains largely unquantified and likely grossly underestimated.

**Fig 1.**
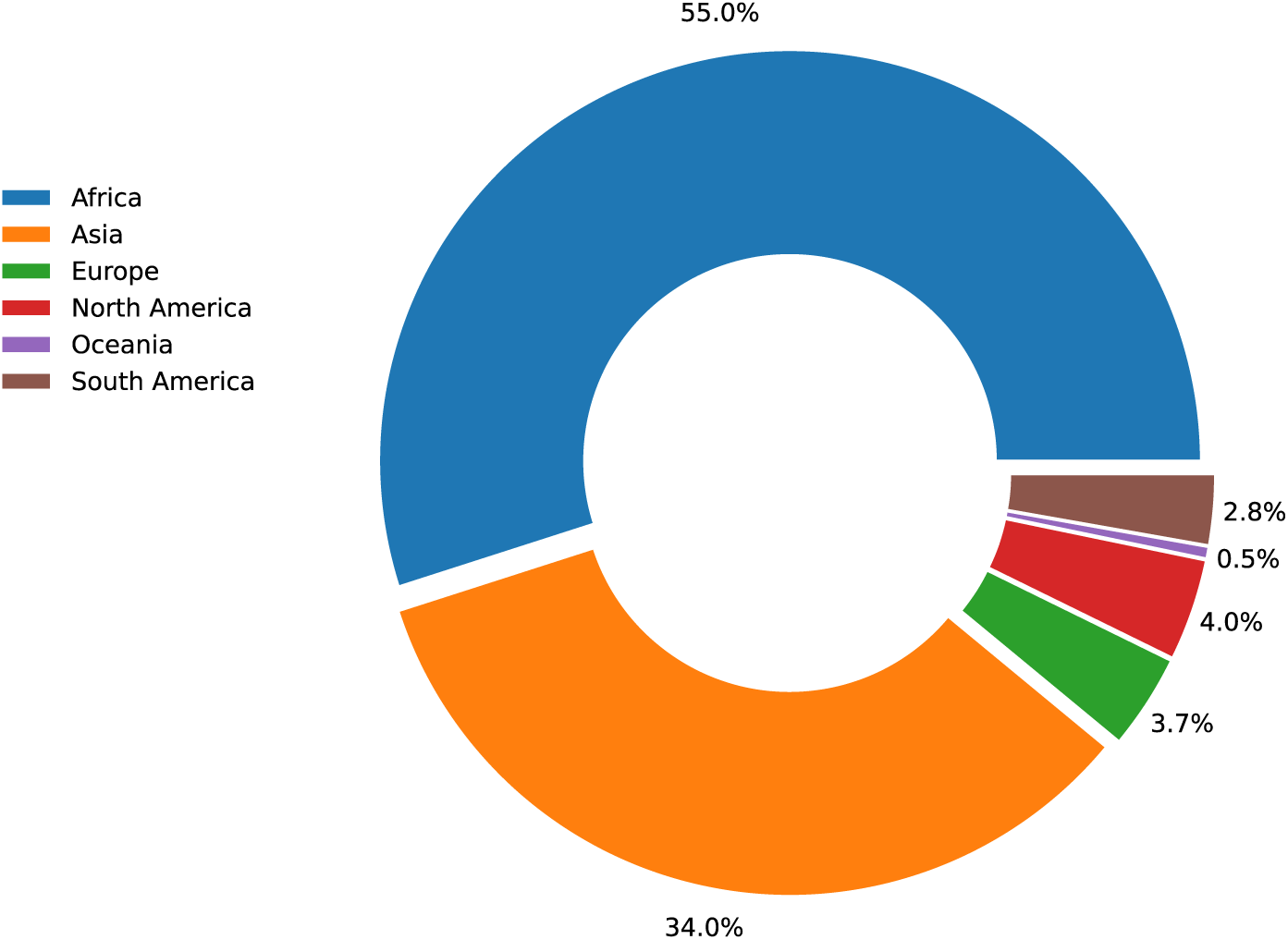
Anthrax outbreaks by region. Worldwide reported animal anthrax outbreaks between 2005 and 2022, shown by continent. There have been 19,171 new outbreaks (shown in the figure) in 106 countries consisting of 226,051 total cases. Data were collected from the World Animal Health Information System (WAHIS) of the World Organisation for Animal Health (WOAH). Last accessed on April 10, 2023.

In order to start addressing major knowledge gaps on the multifaceted costs of NZDs for livestock-dependent populations, we aimed to document the occurrence of anthrax in livestock and humans and to quantify its economic impacts in an endemically affected population of northern Tanzania that is representative of pastoral livestock production systems in rural sub-Saharan Africa. Specifically, we quantified the animal health costs of anthrax on livestock-owning communities and their repercussions for livelihoods. We also assessed human health costs, both in terms of morbidity and mortality. Finally, we determined factors associated with households’ willingness to pay for expenses related to reducing the burden of both human and animal anthrax. Overall, we provide the first comprehensive study of the multiple impacts of anthrax on human health and livelihoods using a multi-faceted approach that could be applied to other NZDs, and identify possible actions for mitigating these impacts.

## Methods

### Study area

The study was carried out in the Ngorongoro Conservation Area (NCA) in northern Tanzania (Fig 2A), which encompasses an area of 8,292 km^2^. It is part of Ngorongoro District, one of seven districts comprising the Arusha region. The NCA is an area with substantial interactions among humans, livestock and wildlife. At the time of the study, the human population of the NCA was estimated at 86,733 [27]. Residents are primarily Maasai pastoralists who have a high dependence on livestock for livelihoods [28]. Dependence on livestock is heightened in the NCA because it is managed as a mixed land use area with high conservation priority, and consequently the cultivation of crops is not permitted. The NCA is a major area for eco-tourism as well as pastoralism, supporting both large wildlife and livestock populations (Fig 2B). Based on a survey of livestock ownership in the NCA, at the time of the study there were approximately 280,000 cattle, 260,000 goats and 325,000 sheep kept based on data available to the authors (CCC, Ngorongoro District Veterinary Officer). There has been some prior evidence to suggest that anthrax is a widespread problem in the NCA [29,30], however, as is the case in most endemic areas, its occurrence in humans and animals and its impact on health and livelihoods had not been quantified prior to this study.

**Fig 2.**
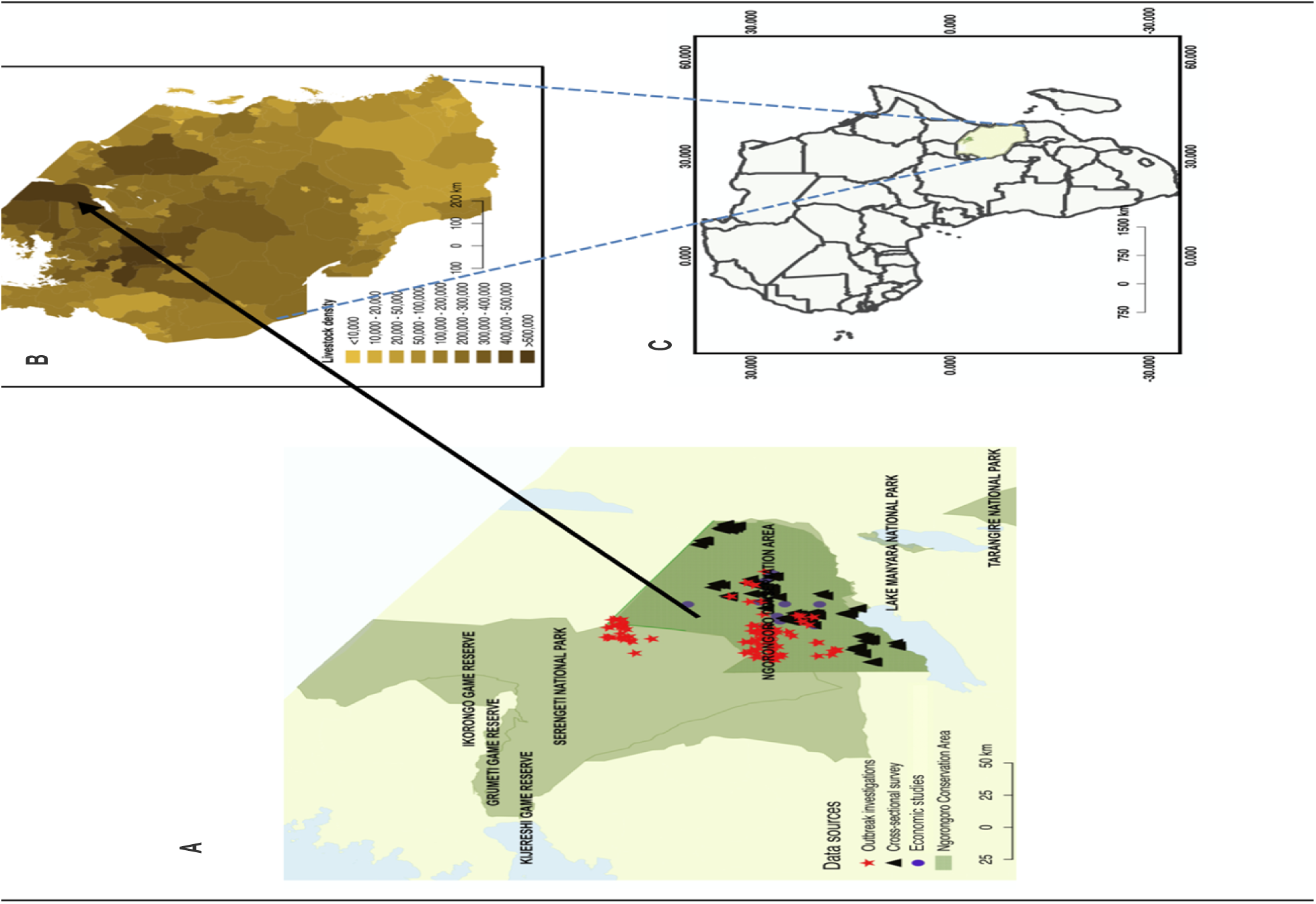
Study area. Maps of the study area in northern Tanzania showing A) the locations within the Ngorongoro Conservation Area from which data generated and analysed in this study originated; B) livestock densities in Tanzania at district level [27]; and C) the location of Tanzania within Africa.

### Data sources

The study was informed by four types of data which are described below.

#### 1. Questionnaire-based household survey

In order to assess the role of livestock in livelihoods, and common causes of morbidity and mortality in livestock, including anthrax, a questionnaire-based household survey was administered between July and October 2016 using a cross-sectional design. The questionnaire was conducted in purposely-selected administrative wards and sub-villages. We chose a representative sample of sub-villages (n = 21) ranging from most to least affected by anthrax (i.e., with different levels of perceived risk) (S1 Text, Appendix A, Figs A-C) based on community knowledge generated through focus group discussions (n = 10). Subsequently, 9 or 10 households were randomly selected from each sub-village, resulting in a total of 209 surveys.

Closed- and open-ended questions were asked about livestock management practices (specifically for cattle, sheep and goats), including causes of mortality, perceived importance of livestock diseases, herd-level morbidity and mortality, and history of anthrax in the household/herd/village with associated losses. More generic questions about household characteristics were also asked, including household and livestock demographics, and livestock use for subsistence and income generation. Participants provided informed consent by writing or thumbprint.

#### 2. Active case detection

In order to capture data on anthrax cases and deaths affecting both animals and humans, active disease surveillance approaches were established. These involved a dedicated team of human and animal health specialists trained in anthrax case recognition and collection of diagnostic samples for case confirmation. The team was responsible for eliciting reporting of cases from study communities, and for investigating and sampling from individual cases/outbreaks.

Cases of animal anthrax (both livestock and wildlife) were actively sought out and investigated between April 2016 and October 2018. Suspected cases were defined as the occurrence of a sudden death in an animal that had previously appeared healthy, or the finding of a carcass showing no signs of predation/starvation/traumatic injuries. Additional signs associated with anthrax, such as blood oozing from the natural orifices and the rapid decomposition of carcasses characterised by swelling [23], were assessed when present. To confirm suspected cases, various samples were collected for testing by quantitative polymerase chain reaction (qPCR) and/or microscopy, namely skin tissue, blood swabs, whole blood and blood smears. Details of sample collection and the diagnostic procedures have been previously outlined [30]. Samples with Ct values <37 for three molecular targets (chromosomal sequence PL3 and plasmid sequences *cap*, located on plasmid pXO2, and *lef*, located on plasmid pXO1) were considered positive, whereas samples were considered ‘suspect’ if they had either a) Ct values < 40 for all targets and at least one target <37; or b) Ct < 37 for at least 2 targets.

For a subset of livestock cases investigated between August 2016 and March 2017, hereby referred to as ‘outbreak investigations’ (n = 54), households were asked additional questions about the number and species of livestock owned, as well as other suspected anthrax cases in their herd, both contemporaneous and historical. These investigations mainly occurred in two wards of the NCA – Endulen (n = 24) and Olbalbal (n = 17); limited investigations were also carried out in Ngoile (n = 8), Esere (n = 3), and Kakesio (n = 2). Confirmation of an anthrax incident among these outbreak investigations depended on it meeting one or both of the following criteria: 1) available sample testing positive for *B. anthracis* either by microscopy or qPCR; or 2) the presence of at least one human pathognomonic anthrax case linked to the outbreak. For this purpose, a human case was defined as a person with a history of possible anthrax exposure (S1 Text, Appendix B) and showing signs and symptoms of cutaneous anthrax. Suspected human cases developing gastrointestinal or inhalational anthrax were not included in the definition, as the symptoms for these forms of the disease are non-specific.

In-depth interviews and physical examinations of suspected human anthrax cases were conducted at Endulen Hospital – the primary health centre for the NCA – and in households between June and December 2018. The objectives of these investigations were to estimate the relative occurrence of the different forms of anthrax (i.e., cutaneous, oropharyngeal - a subtype of cutaneous anthrax occurring within the mouth and/or throat, gastrointestinal, inhalational), and, for those conducted in households, to document cases that did not present at health facilities. The case definitions used were based on those developed by the US Centers for Disease Control and Prevention (CDC) [31] and were adapted to the study area in consultation with local medical experts (S1 Text, Appendix B).

#### 3. Human anthrax case reports from local medical facilities

For human anthrax, active case detection methods were complemented by collection of data on suspected cases reported by health facilities throughout Ngorongoro District (comprising the NCA and the neighbouring area of Loliondo) between March 2017 and October 2019. These data were compiled by the District Medical Office (LK). Additionally, summary annual data on suspected human anthrax cases seen at Endulen Hospital between 2015 and 2017 were compiled by the medical officer in charge (NM), with more detailed case data available for the period from February 2017 to April 2018. This list of reported patients was cross-verified with the cases investigated at the household level during active surveillance to assemble a minimum list of suspected cases for this period. Authors providing the original data had access to information that could identify individual participants, after which anonymised data were shared for analysis.

#### 4. Socio-economic surveys

A cross-sectional survey was carried out between September 2018 and June 2019, inclusive, in order to obtain information on the socio-economic characteristics of households affected by anthrax, and to understand households’ willingness to invest in anthrax mitigation strategies. Regarding the latter, in order to estimate the willingness to pay (WTP) for particular anthrax prevention and control measures, we used choice experiments [32–34]. Specifically, 71 households responded to 48 choice experiments (16 relating to livestock health and 32 relating to human health), resulting in a total sample of 3,408 observations (S1 Text, Figs D-F). The livestock health experiments varied based on four factors: livestock type (cattle vs. small ruminants), disease mitigation options (treatment vs. vaccination), price (high vs. low), and efficacy (highly vs. less efficacious). Similarly, human health choice experiments varied based on five factors: age (adult vs. child - to establish whether people place differing value on their own health compared with that of their children), treatment type (inpatient vs. outpatient), disease type (cutaneous vs. gastrointestinal – the two predominant forms of anthrax in the area), price (high vs. low), and efficacy (highly vs. less efficacious). Each dichotomous choice experiment was followed by an open-ended question about the respondents’ maximum WTP for the given disease mitigation alternative. The prices set were determined by the current local cost of these services and through question pre-testing.

### Data analyses

Data relating to household demographics, including sources of income, levels of education and livestock ownership were summarised (S1 Text, Table A). General mortality of livestock, including deaths suspected or confirmed to be due to anthrax, were quantified. The history of anthrax in households and perceived importance of the disease were described using summary statistics. Logistic regression was used to test whether the probability of a household being affected by animal anthrax was associated with the socio-economic characteristics of the household. These included gender and age of the head of household, income, savings and education.

To conservatively estimate the monetary cost to livelihoods associated with anthrax, direct costs of livestock losses that survey respondents attributed to this disease were calculated. To do this, we summed losses experienced by affected households, using information on livestock prices from the Livestock Information Network Knowledge System (LINKS) for Tanzania (http://www.lmistz.net/Pages/Public/Home.aspx). Where data were available (for current cases in the outbreak investigations), calculations were based on the local prices (based on LINKS) of the species affected. When species information could not be recalled, e.g. when households had lost a large number of animals of different species over a two-year period, the average price of sheep was used. Sheep are considered the species most affected by anthrax in the NCA and are the livestock of lowest value in the area. This ensured that the estimates were conservative rather than overstated. The estimates of losses were computed in United States dollars (USD) using the exchange rate (average between 2015-2016) of 1USD to 2,157.75 Tanzanian shillings (TZS) (https://www.oanda.com/).

We described the impact of anthrax incidents on households’ livelihood and treatment seeking. The median costs (time and financial costs) of treatment for affected household members were calculated. Households’ WTP for anthrax vaccination and treatment were described using summary statistics. Additionally, we used a distribution-free non-parametric Turnbull estimator to predict the lower bound WTP of the respondents, which offers conservative WTP estimates [35] (S1 Text, Appendix C). To analyse the relative importance of the factors included in the choice experiments on the respondents’ WTP, we conducted a Poisson regression of choice counts on the factors included in our choice experiment design (price, efficacy, treatment type, disease type, species considered in the animal health treatment and age in the human health treatment) (S1 Text, Appendix C).

All analyses were carried out in the R statistical program V 3.6.0 using RStudio (RStudio Team, 2018) and Python 3.8.8 using Jupyter Notebook 6.4.8.

### Ethical considerations

Research and ethical approvals were granted by the National Institute for Medical Research (NIMR), Tanzania, with reference number NIMRJHQ/R.8a/Vol. IX/2660; Tanzanian Commission for Science and Technology (COSTECH) number 2016-94-NA-2016-88; Kilimanjaro Christian Medical University College Ethics Review Committee (certificate No. 2050); and the University of Glasgow College of Medical Veterinary and Life Sciences ethics committee (application number 200150152). As part of ethical obligations, informed consent was obtained from community leaders and all participants involved in the study after provision of information regarding the project and its objectives as well as their rights as study subjects. Approved consenting procedures were verbal or written. Any participants under the age of 18 required consent from a parent or guardian. Interviews were carried out with heads of households or, if not possible, other adult household members in their language of choice, Swahili or Maasai. All data collected were analysed anonymously, ensuring the confidentiality of participants.

### Data availability

Data related to this manuscript are available in the University of Glasgow’s Enlighten Research Data repository at https://doi.org/10.5525/gla.researchdata.1870.

## Results

### Role of livestock in livelihoods

Among the 209 households enrolled in the questionnaire based cross-sectional household survey, 207 owned livestock, generally a mix of cattle, sheep and goats (S1 Text, Table B). Other animal species, including donkeys, dogs and chickens, were kept in lower numbers. The main source of income in nearly all households was the sale of livestock or their products (93.3% of respondents; Fig 3A). Livestock serve multiple other purposes in this area. They represent a direct source of food (meat, milk and blood), forming the basis of nourishment (Fig 3B). Hides are used as mattresses and mats for sitting and lying, and to make clothing, shoes and belts; animal fat is used for cooking and making products for personal hygiene such as pomades; and houses are built from hides, mud and animal dung. Apart from livestock use for nutrition, clothing, and shelter, income from the sale of livestock is used for healthcare and educational needs, e.g. paying hospital bills and school fees.

**Fig 3.**
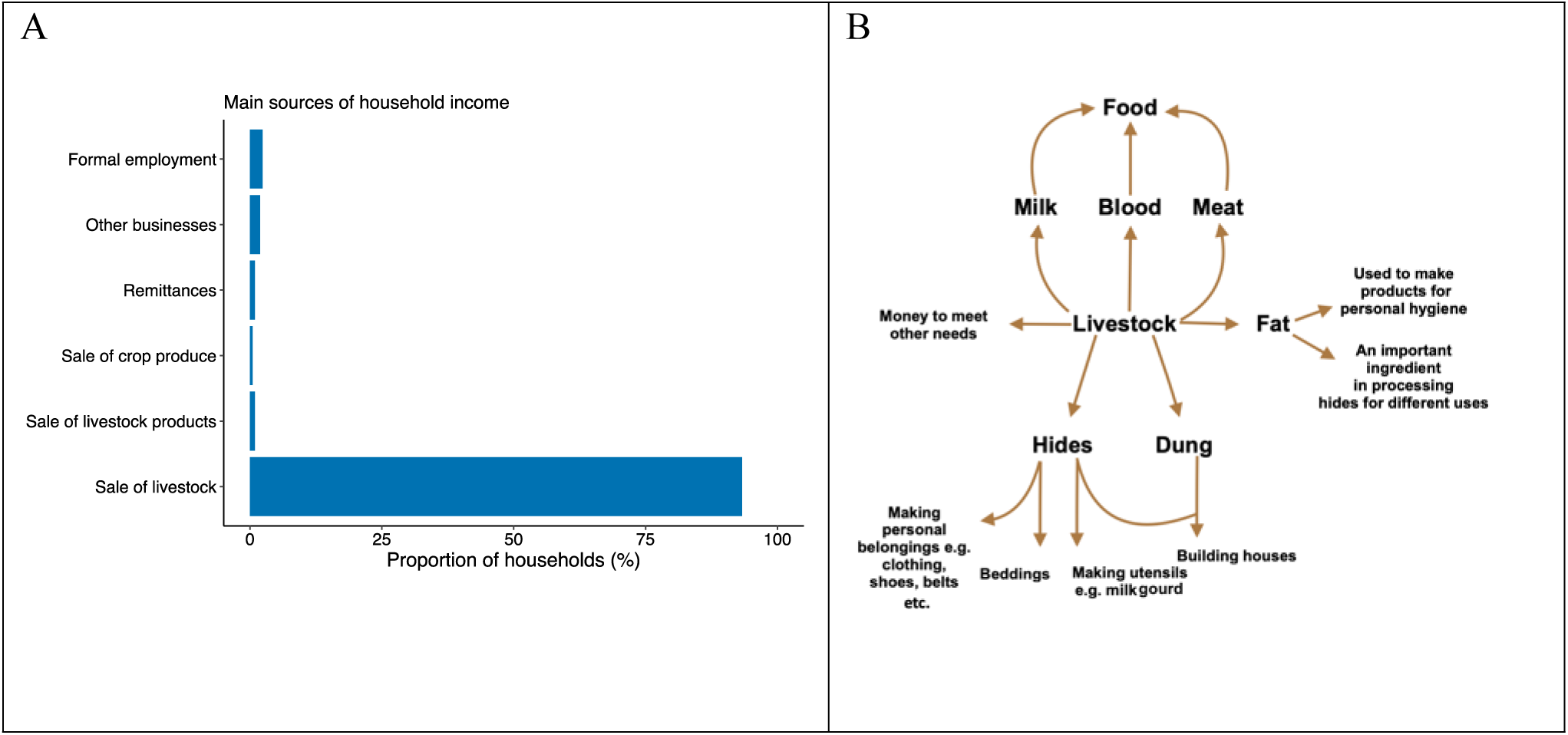
The critical roles of livestock in livelihoods. A) Histogram showing the proportion of households in the Ngorongoro Conservation Area, northern Tanzania, reporting the main sources of income for subsistence, with the sale of livestock representing the primary source. B) Schematic representation of the multiple uses of livestock and their products as reported by study participants. Livestock provide nutrition, shelter, as well as income for other needs like education and health care. Data were obtained through a cross-sectional survey carried out in 209 households between July and October 2016.

### Livelihood impacts of livestock-associated losses

#### Causes of mortality in livestock

Overall, 70.8%, 81.3% and 68.9% of households reported deaths among their cattle, goats and sheep, respectively, in the 12 months preceding the cross-sectional survey. Disease was the most commonly reported cause of these deaths and, in small ruminants, was responsible for higher mortality than drought, predation or other causes (Table 1).

**Table 1.** Perceived causes of mortality in livestock in the Ngorongoro Conservation Area. Data were obtained from a cross-sectional survey conducted in 209 households in 2016, where questions pertained specifically to the 12 months preceding the study.

|  | Cause of mortality |  |  |  |
| --- | --- | --- | --- | --- |
|  | Disease | Drought | Predation | Others<sup>1</sup> |
| Mean number of cattle deaths per household in 12 months (total number of affected households) | 8 (n=119) | 14 (n=50) | 3 (n=33) | 12 (n=7) |
| Mean number of goat deaths per household in 12 months (total number of affected households) | 14 (n=140) | 7 (n=14) | 7 (n=49) | 7 (n=3) |
| Mean number sheep deaths per household in 12 months (total number of affected households) | 15 (n=110) | 4 (n=13) | 6 (n=34) | 3 (n=3) |

<sup>1</sup> From poisoning, flooding and trauma

#### The perceived importance of anthrax for livestock

Among six selected diseases common in livestock in northern Tanzania [36], including anthrax, black quarter, brucellosis, East Coast Fever, foot-and-mouth disease, and Rift Valley fever, nearly half (48.5%) of the surveyed households that responded to this question (n = 134) considered it to be the most important disease for livestock (S1 Text, Fig G). These perceptions varied among administrative wards, with anthrax considered the most important disease in 58.6% of households (41/70) in high-risk areas compared to 31.4% (16/51) of households in low-risk areas (S1 Text, Fig H). As many as 81.0% of respondents living in Olbalbal ranked it as the livestock disease of greatest importance, even when parts of Olbalbal had been classified as low risk areas based on focus group discussions.

### The occurrence of anthrax in animals

Nearly one third (63 of 209, or 30.1%) of the 209 households surveyed reported having lost livestock to anthrax at some point in the past. About 25.0% had experienced anthrax in the 24 months prior to the study, and 20.6% in the 12-month period preceding the study. Reports of disease occurrence were geographically heterogenous but occurred both in areas considered high- and low-risk (S1 Text, Table C). Overall, all households surveyed in Ngoile, 85.7% in Olbalbal and 25.6% in Endulen wards reported past cases of anthrax in their livestock (Fig 4A). In contrast, no households in Eyasi and Ngorongoro wards reported the disease in their livestock, even though human cases were reported by households in the Ngorongoro ward (S1 Text, Appendix A, Fig 4B). We found no statistically significant association between households’ socio-economic characteristics and having a history of anthrax in their livestock (S1 Text, Table D).

**Fig 4.**
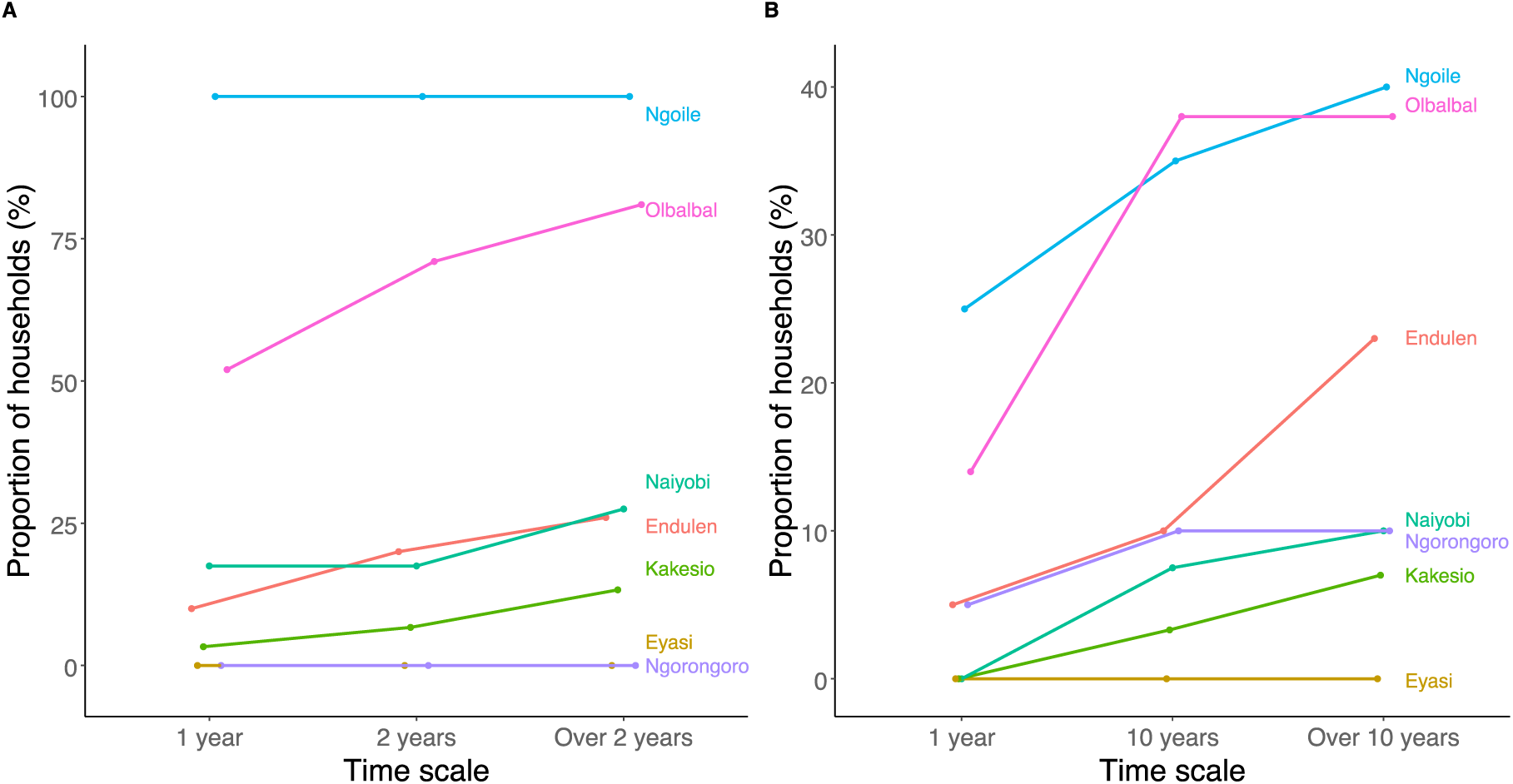
Occurrence of livestock and human anthrax in the Ngorongoro Conservation Area, northern Tanzania. Proportion of households reporting cases of suspected anthrax in their livestock herds (A) and among family members (B), across different timescales in different administrative wards. Data were obtained from a cross-sectional survey of 209 households in 2016. Note that the scales of both axes differ between figures (A) and (B).

Through active case detection, five hundred forty-eight anthrax-suspected animal carcasses reported by community members were sampled and tested between April 2016 and October 2018 (Fig 5; S1 Text, Table E). Of these, 403 (73.5%) were positive by qPCR, and a further 11 samples (2%) gave suspect results, demonstrating that livestock owners were able to very reliably recognise cases. Of the confirmed anthrax-positive cases where the species was recorded (n = 383), the majority were sheep (n = 268), followed by goats (n = 61), cattle (n = 33) and donkeys (n = 6). Several wildlife cases (n = 15) were also confirmed, notably in wildebeest, zebra and elephant. Cases occurred throughout the year, but numbers were heterogeneously distributed, with a clear peak in cases observed between July and November 2017. This corresponded to a particularly severe dry season (July – November) which, based on focus group meetings, is the peak season for anthrax in the NCA.

**Fig 5.**
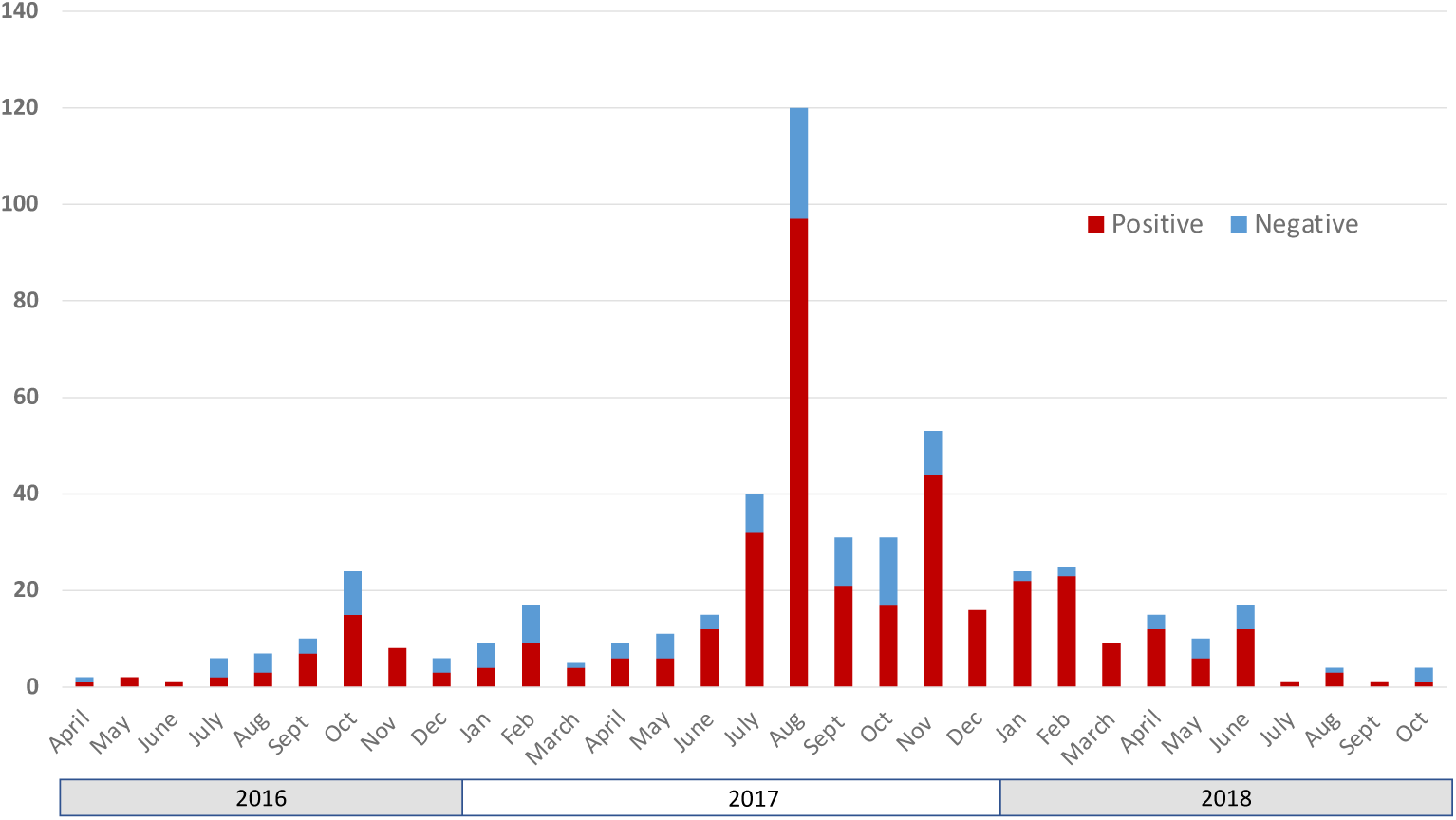
Suspected cases of anthrax in animals in the Ngorongoro Conservation Area, northern Tanzania. Samples were collected from suspect carcasses identified through field surveillance conducted by the research team between April 2016 and October, 2018, and considered positive if confirmed by molecular (qPCR) testing.

### Monetary losses associated with anthrax cases in livestock

Based on responses obtained during household surveys, anthrax occurrence in livestock in particular wards (i.e. Ngoile and Olbalbal) was so commonplace that it was difficult for some households to keep track of the number of animals dying from the disease in a 12- or 24-month period. The estimates reported here are therefore based on information from 55 households, which represents 86% of the households reporting a history of anthrax in their herd. Over the 2-year period, from mid-2014 to mid-2016, households affected by anthrax reported a median number of 10.5 animals lost to suspected anthrax. Moreover, 27.4% of households (15/55) estimated losing over 100 animals and 9.1% (5/55) over 200 animals (S1 Text, Fig IA). Whilst most households lost only one to five animals over the entire 2-year period, households experiencing the highest disease incidence reported up to three suspected cases of anthrax every week during the dry season. In 2015 and 2016, the average price of a sheep was TZS 64,348.42, the equivalent of USD 30. Based on this, the conservative estimated cost of losing livestock to anthrax ranged between USD 30 and USD 6000 per household for the loss of 1 to 200 animals. Thus, for surveyed households recalling at least one suspected case of anthrax in the preceding two years, the estimated asset losses for these 55 households amounted to a combined total of USD 82,094.

Additionally, to obtain more precise estimates of losses through laboratory confirmation, 54 outbreak investigations were conducted between August 2016 and March 2017 as part of the field surveillance implemented for this study. Anthrax was confirmed to be the cause of death for 37 (69.0%) of these incidents. Incident confirmation was based on molecular diagnosis using qPCR (n = 25), microscopy (n = 1), or the occurrence of at least one associated pathognomonic human case (n = 11). A total of 506 livestock were lost in the 37 confirmed outbreaks (Table 2). Sheep were the species most often lost (30 incidents, 411 animals), followed by cattle and goats (S1 Text, Table F). The total number of sheep lost in those incidents was more than 7-fold that of cattle or goats. The median loss experienced by a household was 250 USD per confirmed anthrax incident. The middle 50% of losses (interquartile range) fell between 125 - 509 USD. The maximum loss experienced by a household in a confirmed anthrax incident was 9910 USD. The total losses experienced by households in the confirmed incidents were 28,100 USD (S1 Text, Fig IB). To place the above-mentioned losses into context, of the 82.3% (n = 172) of households willing and able to provide detailed information about monthly household income, the maximum reported monthly income was 400,000 TZS, equivalent to 185 USD which, for a median family size of 9, amounts to 69 cents per person per day.

**Table 2.** The costs of anthrax in livestock in the Ngorongoro Conservation Area. Monetary costs (in USD) of livestock losses associated with 37 confirmed anthrax incidents between August 2016 and March 2017 were calculated (number of animals lost * local unit value in USD for each species). Livestock prices (2015-2016) were based on data from the Livestock Information Network Knowledge System (LINKS) for Tanzania (http://www.lmistz.net/Pages/Public/Home.aspx).

| **Livestock species (number of outbreaks involved)** | **Number of animals lost** | **Median number of livestock lost per outbreak (range)** | **Median proportion of herd lost per outbreak (range)** | **Local unit value (USD)** | **Total value (USD)** |
| --- | --- | --- | --- | --- | --- |
| Cattle (n=9) | 58 | 2 (1-35) | 1.3% (0.2 - 11.7%) | 250 | 15,022 |
| Sheep (n=30) | 411 | 6 (1-80) | 1.6% (0.2 – 28.0%) | 30 | 12,330 |
| Goats (n=7) | 37 | 2 (1-20) | 4% (0.5 - 23.3%) | 31 | 1,147 |

During these investigations, livestock owners described 104 additional incidents of anthrax in their herds within the previous 2 years (excluding the current outbreak), each lasting between a day and up to four months. Among the 158 incidents (both current and past), a total of 2455 livestock (cattle, sheep and goats) were reportedly lost. When adding reported losses in the same herds (with current confirmed cases) during past incidents between 2015 and 2017, the total losses amounted to 88,500 USD (S1 Text, Fig IC; Table G).

Anthrax cases in livestock had several reported impacts on affected households. Those most commonly reported were: 1) an increase in time devoted to livestock care and management (94.7% of respondents); 2) an increase in consumption of livestock that have died from anthrax (78.9%); 3) having to identify new grazing areas or water sources (73.7%); 4) increased monetary costs of livestock management (26.3%); and 5) a decrease in livestock sales (21.1%).

### The occurrence and impacts of anthrax in humans

Anthrax in humans was also common in the NCA, both at the household level and at medical facilities within the area. According to results from questionnaire-based household surveys, one in six households (16.7%) reported having had at least one case of anthrax among family members at some point in the past (Fig 4B). In the 12 months preceding the surveys, 5.7% of households had experienced human anthrax. The proportion of households reporting a history of anthrax in people was as high as 40.0% in certain wards (Fig 4B), with 3.8% of households reporting at least 2 cases and 1% reporting at least 3 cases. Four households interviewed (1.9%) reported the death of a family member due to anthrax.

From 2015 to 2017, between 20 and 89 cases of human anthrax were reported annually by the local medical facility (Endulen Hospital) (S1 Text, Table H), while between 154 and 299 cases were reported annually throughout the Ngorongoro District (NCA and Loliondo) from 2017 to 2019 (S1 Text, Table I; 729 cases in total); these latter figures include data from Endulen hospital. Thirty-four (4.7%) of the cases reported at the district level over this three-year period were fatal. Detailed records were available for a subset of cases reported by the District from 2017 and 2019 (n = 197). Among these patients, 95 were female (48.2%) and 101 were male (51.3%), with sex unreported for one entry. Ages of these patients ranged from 1 to 70 years, with a median age of 15. Eighty-one of these patients (41%) were ten years of age or younger. The majority of cases were cutaneous anthrax, although at least eleven cases of suspected gastro-intestinal anthrax were reported, four of which were fatal.

For the period over which detailed case data were available from both Endulen Hospital and surveillance conducted by our field team (February 2017 - April 2018), at least 33 cases of the 151 observed at the household level (21.9%) did not present at this health facility. The sex ratio of those affected was similar to that observed at the district level with 48.3% of cases being female (n = 73) and 51.6% male (n = 78). Nearly half of all reported cases were among children aged 10 or younger (n = 73), and 29 of these were under 5 years of age.

Fifty-eight anthrax-suspect patients were interviewed and examined in detail by clinical officers in 2018. Of these, 55.1% were female (n =32); eleven patients (19.0%) were under 5 years of age. Fifty-four (93.1%) presented with cutaneous anthrax, three (5.2%) with suspected gastrointestinal anthrax, and one with oropharyngeal anthrax.

Consequences for households experiencing human anthrax included increased expenditure on healthcare (94.7%); an increase in time spent working (89.5%); and increased anxiety (10.5%).

### Determinants of choices related to anthrax mitigation for people and animals

Of the 71 households that responded to the socio-economic survey, nineteen reported having experienced cases of human anthrax. A high proportion (17/19, 89.5%) of affected households sought treatment for the individual(s) affected, although this may be due in part to the assistance provided through the research project, as 35.3% (6/17) mentioned that the availability of clinical personnel on the team influenced their decision to seek treatment. Whenever possible (i.e., if the team was in the field), assistance (transport to hospital, medication) was provided to affected individuals through a clinical officer recruited by the project, in line with the ethical principle of beneficence. For those who sought treatment, the primary factor influencing the decision to seek healthcare, as reported by these households, was the ability to find and fund transport to a healthcare facility (94.1%), followed by the perception of the severity of anthrax (88.2%) and the affordability of treatment (58.8%). Cost of treatment was financed through the sale of an animal in 47.0% of instances, whereas 35.3% obtained free care. Apart from those households that obtained free care, the median cost of seeking care (including travel and treatment costs) for anthrax was 19.6 USD (IQR: 17.1-37.3 USD). In addition to these monetary costs, the median time lost in seeking treatment was 9.0 hours (IQR: 4.3-18.5 hours). In general, access to veterinary services in the study area was variable; households reported travel times ranging from half an hour to six hours in order to access the local livestock field office (S1 Text, Table J).

Characteristics of the respondents to the WTP survey are summarized in S1 Text (Tables J and K). The Poisson regression model outcomes for the choice counts by experiment (price, efficacy, treatment type, disease type, species considered in the animal health treatment and age in the human health treatment) are presented in S1 Text, Table L.

Willingness to pay (i.e., positive response rates) for 16 animal health choice experiments are presented in Fig 6. Respondents tended to reject disease mitigation strategies with low (50%) efficacy, which were not included in the following analysis. Even with high (100%) efficacy, responses varied significantly between low-price and high-price mitigation options, implying high price sensitivity. A joint test using Cochran’s Q test rejects the null hypothesis that the response rates in the 8 experiments are equal at the 1% significance level (Cochran’s Q = 206.8, df = 7, p-value = 0.00) [37]. For low-price mitigation options, the positive response rates for WTP ranged between 73% and 86%, while for high-price alternatives, the positive response rates were between 14% and 24%. The pairwise comparisons of the 8 experiments regarding mitigation alternatives that varied by price only (low vs high), while holding species and treatment type fixed, were significant at the 1% level using Cochran’s Q tests. Within the four low-price disease mitigation alternatives, there was a statistically significant difference in responses at the 10% level (Cochran’s Q = 6.59, df = 3, p-value = 0.086). Further disaggregating by species within low-price alternatives, we found significant differences across treatment and vaccination for cattle only. The difference between treatment and vaccination was significant in cattle disease mitigation at the 10% level (Cochran’s Q = 3.27, p-value = 0.07), but not significant in small ruminants.

**Fig 6.**
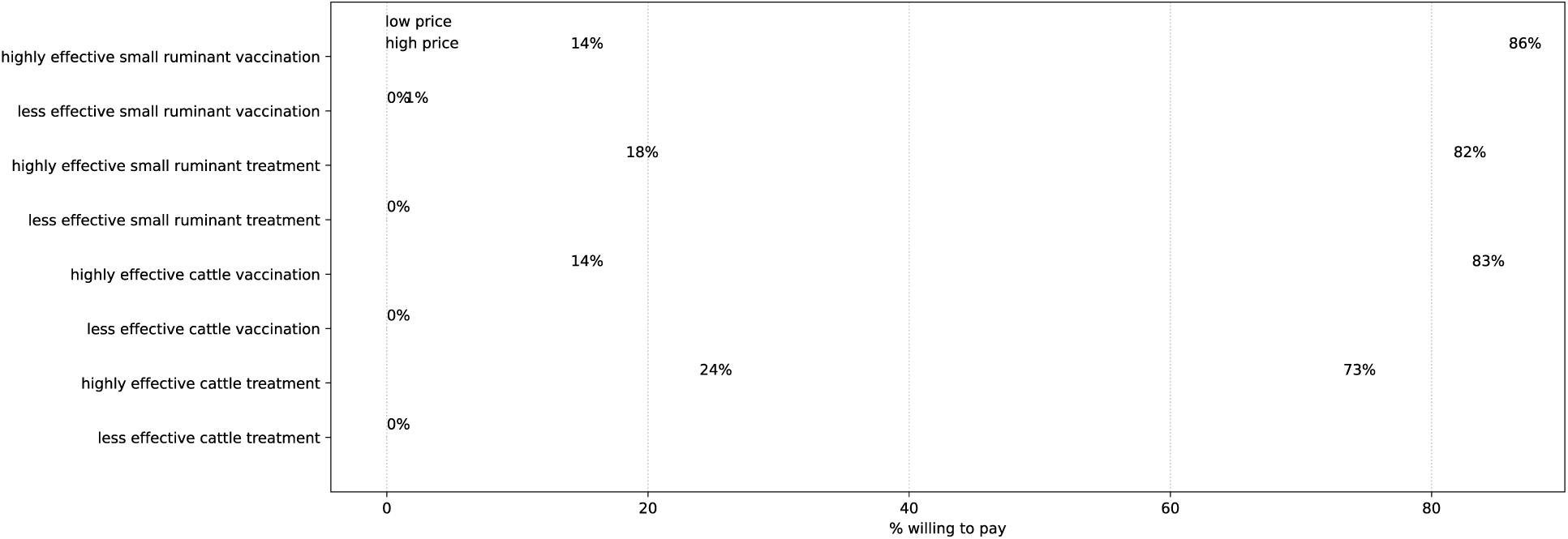
Frequency of positive responses in animal health experiments. Blue and green dots show response rates in low-price and high-price disease mitigation choices, respectively. The dashed lines indicate the difference in willingness to pay for low-price and high-price options in each disease mitigation alternative considered. Differences are considered between highly (100%) effective and less (50%) effective interventions.

Within the four high-price alternatives, overall response rates did not differ (Cochran’s Q = 5.35, df = 3, p-value = 0.15). While disaggregated by species, the treatment and vaccination adoption differed in cattle disease mitigation at the 10% level (Cochran’s Q = 3.77, p-value = 0.052), but not in small ruminants.

Scenarios in which respondents were willing to pay for human health interventions are summarized in Fig 7 for 32 different choice experiments. Similar to the animal health choice experiments, the responses for human health interventions exhibited price sensitivity (all pairwise comparisons were significant at the 1% level) with complete reluctance to adopt any treatment options with low (50%) efficacy, which were not included in the following analysis. There was a noticeable difference in the treatment uptake rate depending on whether the treatment was received by adults or children. In the eight low-price treatment mitigation options, the response rates were statistically different at the 10% significance level (Cochran’s Q = 13.52, df = 7, p-value = 0.06). Within these low-price options, pairwise comparison by patient age (adult vs child) with fixed disease and treatment type showed a difference in the positive response rates between adults and children for inpatient treatment being significant at the 5% level (cutaneous anthrax: Cochran’s Q = 4.84, p-value = 0.03; gastro anthrax: Cochran’s Q = 4.17, p-value = 0.04).

**Fig 7.**
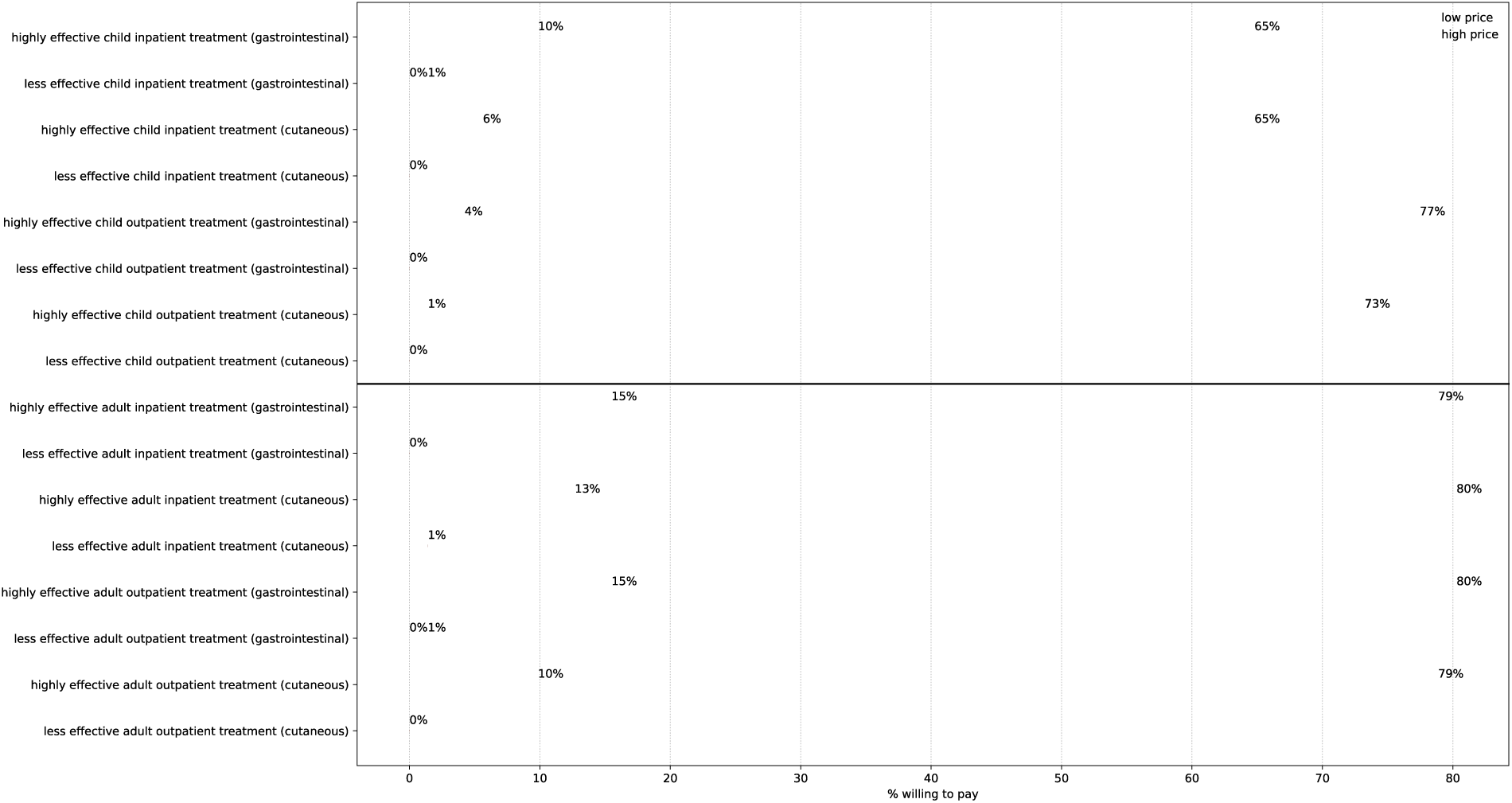
Frequency of positive responses in human health experiments. The top panel shows the positive response rates for 16 choice experiments regarding treatments for children, and the bottom panel shows those for adult treatments. Blue and green dots show response rates in low-price and high-price disease mitigation choices, respectively. The dashed lines indicate the difference in willingness to pay for low-price and high-price options in each disease mitigation alternative. Differences are considered between highly (100%) effective and less (50%) effective interventions.

In the eight high-price treatment mitigation options, the response rates were statistically different at the 1% significance level (Cochran’s Q = 24.97, df = 7, p-value = 0.00). Pairwise comparison by patient age (adult vs child) of these options showed the positive response rates differed significantly at the 5% level between adults and children for all treatment options except inpatient gastrointestinal anthrax treatment.

The overall response rates revealed the importance of the relative price of treatment options in household resource allocation. Exceptions arose when the relative monetary price was too high, such as treatment for inpatient gastrointestinal anthrax in high-price options (Tsh. 60,000 for adults, Tsh. 40,000 for children) or the relative price was too low, such as outpatient treatment in low-price options (price ranging from Tsh. 10,000 to Tsh. 20,000), wherein the household choices do not vary by patient age.

Fixed price bids for each choice experiment presented to the respondents, along with the average of the stated maximum WTP (presented as an open-ended question after the fixed price bids) are reported in Table 3. The stated maximum WTP of the households was persistently close to the value of the lower bids in animal health experiments, whereas for the human health experiments, the stated maximum WTP was always below the lower bids. Respondents stated a willingness to pay more for the treatment of human vs. animal anthrax. Human health mitigation options incur considerably higher monetary costs. Table 3 (last two columns) also reports lower-bound WTP estimates using the Turnbull estimator and their 95% confidence intervals, which offers conservative estimates of WTP. The estimated (Turnbull) WTP are often below the stated maximum WTP, as expected, implying the respondents’ reluctance to adopt high-cost alternatives. However, the estimated WTP in the adult’s health experiments often exceeded the estimated amount in children’s treatment alternatives, which is consistent with the stated WTP, reinforcing our findings of lower propensity to pay for the children’s anthrax treatment.

**Table 3.** Willingness To Pay (WTP) bids, stated and estimated WTP by experiment in the survey. Columns 2 and 3 report the fixed low-price and high-price bids (in Tanzanian shillings) used in the choice experiments. Column 4 reports the mean stated maximum WTP amounts by respondents. The last two columns show the estimated lower-bound WTP and their confidence intervals using the Turnbull estimator, which offers a conservative WTP estimate (see details in S1 Text, Appendix C). Only the choice experiments with high efficacy are reported.

| **Experiment** | **Low bid** | **High bid** | **Mean of Stated Max WTP** | **Lower-bound WTP (Turnbull)** | **95% CI (Turnbull)** |
| --- | --- | --- | --- | --- | --- |
| Cattle Treatment | 3000 | 6000 | 3084 | 1479 | [1050; 1908] |
| Cattle Vaccination | 400 | 800 | 188 | 276 | [228; 324] |
| Small Ruminant Treatment | 200 | 400 | 223 | 127 | [101; 152] |
| Small Ruminant Vaccination | 200 | 400 | 201 | 144 | [121; 167] |
| Cutaneous Outpatient Treatment for Adults | 15,000 | 30,000 | 11,950 | 10,352 | [8588; 12,116] |
| Cutaneous Inpatient Treatment for Adults | 25,000 | 50,000 | 22,795 | 12,958 | [10,456; 15,460] |
| Gastrointestinal Outpatient Treatment for Adults | 20,000 | 40,000 | 18,676 | 16,901 | [13,885; 19,917] |
| Gastrointestinal Inpatient Treatment for Adults | 40,000 | 60,000 | 33,605 | 25,352 | [21,198; 29,507] |
| Cutaneous Outpatient Treatment for Children | 15,000 | 30,000 | 5908 | 10,775 | [9176; 12,373] |
| Cutaneous Inpatient Treatment for Children | 15,000 | 30,000 | 7605 | 7324 | [6245; 8403] |
| Gastrointestinal Outpatient Treatment for Children | 10,000 | 20,000 | 5788 | 8873 | [7023; 10,724] |
| Gastrointestinal Inpatient Treatment for Children | 20,000 | 40,000 | 12,492 | 10,986 | [8367; 13,605] |

Table 4 presents the mean stated WTP for children’s treatment by different demographic factors, which exhibits marginal differences in the WTP for treatment of children depended on household size, parent gender, and respondents’ experience. Specifically, larger households and male respondents, on average, stated lower WTP amounts. In addition, older respondents and those with recent experience tended to state a lower WTP amount.

**Table 4.** Mean stated willingness to pay (WTP) for anthrax treatment of children by different respondent characteristics. Respondents’ stated WTP for treatment options for children is decomposed by their recent experience with anthrax, household (HH) size, sex and age.

|  | HH size |  | Sex of household respondent |  | Age of respondent |  |  | Recent experience |  |
| --- | --- | --- | --- | --- | --- | --- | --- | --- | --- |
|  | Below average | Above average | F | M | 18-35 | 36-59 | > 59 | Yes | No |
| Child inpatient treatment for cutaneous anthrax | 8833 | 6343 | 9240 | 6717 | 7609 | 7805 | 6429 | 7167 | 7830 |
| Child inpatient treatment for gastrointestinal anthrax | 13611 | 11343 | 13000 | 12217 | 12913 | 13049 | 7857 | 10625 | 13447 |
| Child outpatient treatment for cutaneous anthrax | 6278 | 5529 | 6120 | 5793 | 7261 | 5768 | 2286 | 4708 | 6521 |
| Child outpatient treatment for gastrointestinal anthrax | 6250 | 5314 | 7280 | 4978 | 7348 | 5488 | 2429 | 5792 | 5787 |

## Discussion

A defining feature of NZDs is a paucity of data on their occurrence and impacts, and action towards filling these knowledge gaps has remained limited. In this study, we focus on anthrax and provide evidence of the high burden and multi-faceted impacts of this zoonosis on vulnerable communities living in endemic areas. We show that anthrax is a persistent problem, causing substantial livestock mortality: most households in the highest risk areas experience regular animal losses, and, in households that experienced the highest number of cases, annual financial losses due to livestock deaths caused by anthrax are nearly five times the average annual household income. Cases of human anthrax in the study area were commonplace, with multiple fatalities reported annually. The occurrence of disease forces households to change livestock management practices, for example having to identify new grazing and watering areas. This can be difficult and time-consuming, particularly where access to these resources is limited. Households also report negative impacts associated with decreases in sales of livestock and their products, healthcare costs and anxiety. Both for human and animal anthrax, households were only willing to contribute to disease mitigation measures with high efficacy and low cost. Overall, our study highlights the substantial but unrecognised impacts of a zoonosis such as anthrax on the health and socio-economic wellbeing of affected communities.

Our study confirms the importance of livestock health and productivity for pastoral communities [38,39]. Nearly every household we surveyed possessed livestock (99%), which provide the basic needs of food, shelter, clothing, etc. (Fig 3B). Additionally, they represent the primary source of income in the NCA (Fig 3A), with livestock sales helping to pay for necessities like healthcare and education. This reliance on livestock means that negative impacts of disease on the health and productivity of these animals are likely to have far-reaching consequences for livelihoods in pastoralist communities, well beyond food security. For example, a study in Kenya showed that protecting the health of livestock by vaccinating them against East Coast Fever improved family income and gender equity, and enabled access to education for girls [40].

Disease was reported as a major cause of livestock deaths – and the main cause for small ruminants – in the households we surveyed, and a high proportion of households ranked anthrax as being the most important livestock disease. Certain communities were disproportionately affected, with more than 85% of households in Ngoile and Olbalbal reporting cases of anthrax among their livestock. This phenomenon has also been observed in Kenya, where particular localities experienced repeated outbreaks over time [41], consistent with the long-term environmental survival of anthrax spores after outbreaks. We confirmed that at least three quarters of all sudden livestock deaths reported by community members were due to anthrax, demonstrating a high level of local disease awareness and strengthening our confidence in survey responses related to anthrax-associated losses. A total of 368 livestock cases tested positive for anthrax over 31 months of active surveillance, which averages to just under 150 cases a year. Our results demonstrate a high degree of under-reporting of anthrax in livestock in Tanzania: based on the national reporting system, a total of 118 livestock deaths (87 bovine, 23 caprine and 8 ovine) were documented for the entire Arusha region (37,576 km^2^) – including the NCA (8,292 km^2^) – between 2013 and 2016 [42]. Even in our study, where reporting was increased due to active case finding, we will have underestimated the true number of livestock cases since 1) our field team could not attend to all reported cases due to limited time and the remoteness of many households; and 2) only one case was often tested per household, despite multiple cases having occurred. Moreover, we expect that not all suspected cases were reported to our field team, particularly from the wards that received less research attention during outbreak investigations. Although the NCA may be a particularly high-risk location for anthrax [29], based on the high number of cases confirmed in this relatively small area and the degree of underreporting observed, it is reasonable to expect that thousands of cases go undocumented each year in Tanzania. Substantial livestock mortality associated with anthrax, with herd-level mortality as high as 28%, has also been reported from Zambia and Zimbabwe [14,15], suggesting that such losses are widespread in eastern Africa.

Livestock losses due to anthrax are staggering in the context of the household incomes in the NCA. Our estimates indicate that average annual losses are around 300 USD per anthrax-affected household, which is equivalent to over a month’s income for the majority of households in the study area. However, due to limitations in data collection and study design, these estimates should be regarded as a guide, not as an exact measurement of the losses attributable to animal anthrax deaths. Nonetheless, not only are these losses significant in comparison to household income, there are much wider benefits of keeping livestock for pastoralist subsistence (e.g., Fig 3B). Thus, the traditional value of livestock, beyond their market value, is important to consider, when measuring the impacts of livestock losses on farmers’ economic and emotional wellbeing [17].

Anthrax was also commonplace in the human population, and we found that four out of five human cases were linked to suspected or confirmed livestock cases. The high proportion of affected households reported in the surveys (one in six), along with the high number of human cases reported at the district level from 2017-2019 and high case fatality ratio (4.7%) – more than 10-fold that of malaria (https://www.who.int/data/gho/indicator-metadata-registry/imr-details/16) – suggests that anthrax continues to place a major burden on public health in this area. High numbers of human cases with comparable case fatality have also been reported in Zambia and Zimbabwe [15,43–45]. Most cases are likely associated with the preparation and consumption of meat from anthrax-suspected carcasses, a practice that remains commonplace in the NCA [24], and is apparently widespread in eastern Africa [46–48]. Indeed, our respondents reported regular consumption of suspected anthrax-infected animals, which highlights the major health risks associated with cases in livestock, as well as the complexities of risk perception in endemic areas.

We observed an increase in the number of cases reported by Endulen Hospital between 2015 and 2017 (Table H), as well as at the district level between 2017 and 2019 (Table I). Similarly, Mwakapeje *et al*. noted an increasing trend in the number of human anthrax cases reported in both the Arusha and Kilimanjaro regions since 2014 [42]. While this could potentially reflect increased anthrax incidence in the area, it is likely attributable at least in part to improvements in human health reporting and research activities; anthrax has been included as an immediately reportable disease in the electronic reporting system (eIDSR) since 2013 [42]. A substantial proportion of anthrax cases in the community (∼22%) do not seek formal medical attention due to limited access, so even with improved reporting based on cases seen at health facilities, the overall burden of anthrax in the human population is underestimated. While the majority of these cases would be expected to be cutaneous anthrax, a form that is largely self-limiting, failure to rapidly seek treatment means that certain cases that do develop more serious complications can result in fatalities.

Access to both human and veterinary healthcare in sub-Saharan Africa is a persistent problem, particularly in the most remote locations [49–51]. Our findings indicated that human anthrax was associated with high costs and time demands associated with the need to seek healthcare, and that the ability to find and fund transportation to a health facility was a more important factor in seeking treatment for anthrax than the cost of the treatment itself. The problem of accessibility is further complicated for pastoral communities who practise transhumance, i.e. seasonally moving livestock to access pasture, water and minerals [52]. The inability of the most remote communities to physically access healthcare facilities thus likely contributes to severe and even fatal outcomes when people contract anthrax. Access to animal health care and professionals is equally problematic in Africa due to underfunding and understaffing of veterinary healthcare systems [53], which is a particular problem in the most remote areas.

Households’ WTP for different anthrax mitigation alternatives was highly sensitive to price and efficacy; participants were reluctant to contribute to measures that had only 50% efficacy or 100% efficacy but high cost. This reluctance might reflect local experience with suboptimal treatment and/or vaccination, both in terms of quality and delivery [54,55]. Those with recent experience with anthrax stated a lower willingness to invest in anthrax prevention or treatment. It would be valuable to follow up to understand the reasons underlying this difference, e.g., whether this is consistent with concerns related to the efficacy of available treatment and prevention options, whether those having experienced less severe disease symptoms might have different perceptions of anthrax risk, or whether this was due to already-depleted household resources from having experienced a recent case.

Households are faced with difficult choices related to expenditures; it appears that when there is greater uncertainty in treatment outcomes, there is significantly lower uptake. The decision to invest in treatment is likely influenced by the severity of the condition; households’ stronger preference and higher WTP for the treatment of gastrointestinal anthrax, which has a higher fatality rate compared to more common cutaneous anthrax, supports this argument. Despite the higher monetary and cultural value placed on cattle in the NCA, surprisingly, there was a higher stated propensity to pay for treatment and vaccination for small ruminants. While the motivation for these choices was not recorded, a possible explanation could be the particular resilience of small ruminants to drought [56], which, given that the most anthrax-affected areas of the NCA are particularly arid [57], makes these species especially valuable for these communities. This could also be associated with the lower monetary costs related to treatment and vaccination of small ruminants, which could be perceived as being more affordable, or the fact that sheep are the species most commonly affected by anthrax in the NCA. Price sensitivity to health choice alternatives is common and supported by previous studies [58–60].

We found WTP for adult treatment of anthrax was significantly higher than that for children. Commonly, we expect an altruistic behaviour among adults with their own health risks compared to those of their children [61–63]. This finding could again reflect the difficult economic choices faced by rural livestock owners. For example, with limited budgets, households may have to choose to allocate resources between the treatment of an income-earning adult or a child based on the net marginal benefits, favouring treating the adult. Further, lower WTP in larger households appears to support the fact that difficult choices must be made when resources are stretched. Anecdotal comments reported from the survey suggest there are expectations that treatment of anthrax in children would or should be subsidised.

There are certain limitations to some of our estimates. Some differences exist between reported animal losses estimated from the surveys and those obtained as part of case investigations. For instance, more households reported the death of a larger number of animals in the surveys compared to the incidence investigations. Part of this discrepancy could be due to recall bias in reporting of historical deaths. Over longer time periods, households could be more likely to remember outbreaks in which a larger number of animals are affected, with potentially lower recall for smaller numbers of intermittent cases. For the estimates obtained through confirmed recent incidents, recall bias is much less likely, thus making it a more reliable way to capture data from both large and smaller outbreaks.

### Conclusions

National and international decisions related to the prioritization and control of infectious diseases depend on the availability of data quantifying the importance and impact a disease has on society. Our study provides strong evidence for the significant and multi-faceted impacts of anthrax in an endemic area, which has been lacking from the scientific literature. Anthrax remains an important neglected zoonotic disease for animal and human health, with significant impacts on livelihoods in livestock-dependent communities. Improving the quality of healthcare in rural areas is an essential first step to limit the losses due to this and other neglected diseases. Research into the efficacy of large-scale livestock vaccination campaigns would be valuable to consider for the control of anthrax in endemic areas, in conjunction with improved disease surveillance [30], multi-sectoral response to outbreaks [64], and community awareness messaging. Subsidising disease mitigation services and payment programs that assist economically vulnerable communities might be necessary for higher participation and success of the disease control strategies, and would help alleviate difficult choices related to health-related expenditure within households. For these approaches to be effective, it will be essential to work with communities to co-develop solutions that would be relevant and feasible for them. As with many neglected zoonoses, optimal strategies will need to involve the joint delivery of human and animal health services.

## Data Availability

Data related to this manuscript are available in the University of Glasgow’s Enlighten Research Data repository at https://doi.org/10.5525/gla.researchdata.1870 or in the accompanying supplementary material.

## Acknowledgements

We are grateful for all the support received for this research, including from the NCA community. We thank the Ngorongoro District Council, Ngorongoro Conservation Area Authority, Tanzania Wildlife Research Institute, and members of our field team - Sironga Nanjicho, Kadogo Lerimba, Paulo Makutian, Mary Lonyori, Christopher Kiboya, and Godwin Mshumba. We are also grateful to the Directorate of Veterinary Services, Ministry of Agriculture, Livestock and Fisheries, and Ministry of Health, Community Development, Gender, Elderly and Children for their support. We thank Suzanna Lewis and Andrew Simpson of Public Health England, Porton Down for helping facilitate training sessions for human health professionals, and Amref Health Africa for contributing to knowledge exchange workshops.

## Funding

ORA was supported by a grant from the Gates Foundation (Program for Enhancing the Health and Productivity of Livestock, project reference ID 1083453). TLF was supported by a Marie Skłodowska-Curie Individual Fellowship (659223), a fellowship from the Natural Sciences and Engineering Research Council of Canada (PDF-471750-2015), and a Biotechnology and Biological Sciences Research Council (BBSRC) Discovery Fellowship (BB/R012075/1). The work was also supported by the Wellcome Trust through a Springboard award to TL by the Academy of Medical Sciences. MMR and TLM received partial support for analysis from the Paul G. Allen School for Global Health, College of Veterinarian Medicine and the School of Economic Sciences, College of Agricultural, Natural Resources, and Human Sciences, Washington State University. MPR received support from the US National Institute of Allergy & Infectious Diseases (K23 AI 116869).

## Authors’ contributions

Conceptualisation: ORA, TLF, RNZ, RB, TL. Data curation: ORA, TLF, CC, LK, NM, DM, SOM, SP. Formal analysis: ORA, TLF, MMR, TLM, SP. Funding acquisition: ORA, TLF, TL. Investigation: ORA, TLF, DM, SOM, TL. Methodology: ORA, TLF, MMR, RNZ, RB, TLM, TL. Project administration: ORA, TLF, BTM, TL. Resources: LK, NM, BTM. Supervision: TLF, RNZ, RB, TL, MPR. Writing – original draft: ORA, TLF, MMR, TLM, TL. Writing – review & editing: TLF, TL, MMR, TLM, MPR, RB, RNZ, ORA.

## Notes

### Competing Interest Statement

The authors have declared no competing interest.

### Author Declarations

Research and ethical approvals were granted by the National Institute for Medical Research (NIMR), Tanzania, with reference number NIMRJHQ/R.8a/Vol. IX/2660 Tanzanian Commission for Science and Technology (COSTECH) number 2016-94-NA-2016-88 Kilimanjaro Christian Medical University College Ethics Review Committee (certificate No. 2050) and the University of Glasgow College of Medical Veterinary and Life Sciences ethics committee (application number 200150152). As part of ethical obligations, informed consent was obtained from community leaders and all participants involved in the study after provision of information regarding the project and its objectives as well as their rights as study subjects. Approved consenting procedures were verbal or written. Any participants under the age of 18 required consent from a parent or guardian. Interviews were carried out with heads of households or, if not possible, other adult household members in their language of choice, Swahili or Maasai. All data collected were analysed anonymously, ensuring the confidentiality of participants.

